# Impact of prescription-free access to sexually transmitted infection screening tests in medical-biological laboratories: cross-sectional analysis of data from clinical laboratories in France

**DOI:** 10.64898/2026.04.23.26351562

**Authors:** Andres Gil-Salcedo, Vincent Gazzano, Stephanie Arsene, Agnes Durand, Steven Roger, Laurence Prots, Nathalie Laurencin, Emmanuel Chanard, Alexis Duez, Erwan Le Naour, Olivier Bausset, Benoist Ghali, Anne-Claire Strzelecki, Claire Felloni, Romuald Levillain, Cecile Fargeat, Sebastien Lefrancois, Delphine Feuerstein, Benoit Visseaux, Laurent Escudie, Claire Visseaux, Charles Leclerc, Stéphanie Haïm-Boukobza, the Cerballiance STI study group

**Affiliations:** Cerba Healthcare, Issy-les-Moulineaux, France; Cerballiance, Auvergne-Rhône Alpes, CerbaHealthcare, France; Cerballiance, Normandie, CerbaHealthCare, France; Cerballiance, Ile-de-France Paris, CerbaHealthCare, France; Cerballiance, Rennes, CerbaHealthCare, France; Cerballiance, Provence Azur, CerbaHealthCare, France; Cerballiance, Nouvelle Aquitaine, CerbaHealthCare, France; Cerballiance, Centre-Ouest, CerbaHealthcare, France; Cerballiance, Occitanie, CerbaHealthcare, France; Cerballiance, Hauts de France, CerbaHealthCare, France; Cerballiance, Centre Val de Loire, CerbaHealthcare, France; Laboratoire Cerba, Frépillon, CerbaHealthcare, France

**Keywords:** Sexually transmitted infection, *Chlamydia trachomatis*, *Neisseria gonorrhoeae*, HIV, Hepatitis B, Syphilis, screening program, laboratories

## Abstract

**Background:** Since September 2024, France has implemented a national reform allowing prescription-free access (PFA) to sexually transmitted infection (STI) screening in medical biological laboratories (MBLs). This study aims to characterize the populations undergoing STI testing according to their access modality and evaluate the probability of test positivity in relation to testing pathway, sex, and age groups.

**Methods:** We conducted a cross-sectional analysis of all individuals screened for *Chlamydia trachomatis*, Gonorrhoea, human immunodeficiency virus (HIV), hepatitis B virus (HBV), and syphilis by treponemal-specific immunoassay (TSI) in Cerballiance MBLs between Mars 2025 and February 2026. Multivariable logistic regression models stratified by sex and adjusted for age and region assessed associations between screening modality and STI positivity.

**Results:** Among 1,008,737 individuals included, 27.8% were under PFA and 72.2 under prescription-based access (PBA). PFA users were more frequently male (47.4% vs. 36.3%, p<0.001) and aged 20-39 years (34.0%, p<0.001). Overall positivity rates differed by modality: PFA was associated with higher detection of *Chlamydia* (4.6% vs. 3.6%). PBA group showed more positive cases of syphilis (3.4% vs. 1.2%), HBV (1.3% vs. 0.4%), and HIV infections (0.3% vs. 0.2%, all p<0.001). Co-infection and gonorrhoea proportions did not significantly differ between modalities.

**Conclusions:** PFA substantially increased STI screening uptake, particularly among young adults and men, and enhanced detection of bacterial STIs. PBA remains essential for diagnosing viral and chronic infections. These findings highlight the complementary roles of both access strategies and support PFA screening as an effective public health intervention to broaden STI detection and reduce transmission.

## INTRODUCTION

Sexually transmitted infections (STIs) remain a major and persistent public health issue worldwide^1^. Global estimates indicate that nearly one million new cases of curable STIs (primarily *Chlamydia*, gonorrhea, syphilis and trichomoniasis) occur each day^2,3^. In addition, substantial disease burdens arise from viral infections such as human immunodeficiency virus (HIV) and hepatitis B (HBV), which lead to chronic disease and premature death^1,3,4^. The individual and societal consequences of untreated STIs are substantial: bacterial infections such as *Chlamydia* trachomatis and *Neisseria gonorrhoeae* may ascend the genital tract and cause pelvic inflammatory disease^5,6^, tubal-factor infertility, ectopic pregnancy and chronic pelvic pain in women^7,8^, and epididymitis in men; syphilis can progress to severe neurological and cardiovascular disease^9,10^; last, chronic HBV infection is a leading cause of cirrhosis and hepatocellular carcinoma^11,12^.

Effective STIs control relies on rapid detection, prompt treatment and interruption of subsequent transmission (classical Test-and-Treat public health paradigm)^13–15^. This strategy aims to shorten the infectious period and reduce community spread by initiating therapy immediately upon diagnosis. Diagnostic methods with high sensitivity and specificity, especially nucleic acid amplification tests (NAATs), provide reliable screening of asymptomatic individuals and have markedly advanced laboratory case-finding capabilities^16^. Screening is recommended across many settings for populations at elevated risk, because identifying asymptomatic infections is key to prevent disease-related complications and to curb onward transmission at the population level^17^. However, the availability of accurate diagnostic tests alone is insufficient; effective STIs control also requires ensuring timely access to testing services and minimizing attrition along the diagnostic and care pathway, so that individuals do not disengage before a definitive diagnosis and appropriate management are achieved.

Several barriers have been identified that limit the effectiveness of screening at the structural, healthcare-system, and individual levels^18^. Common barriers include mandatory medical consultations, prescriptions requirement, restricted office hours, and fragmented or complex referral pathways^18–20^. In addition to the aforementioned factors, practical constraints such as needing time off work, transportation difficulties, and the requirement for multiple visits further contribute to reduced uptake and completion of screening^18,20–23^.

These obstacles are of particular significance for groups with limited routine engagement with healthcare services, such as adolescents and young adults, even when their incidence rates exceed those of other groups^18,21,23^. As a result, the cumulative impact of these barriers leads to attrition across the screening continuum and undermines population-level control efforts aimed at identifying underdiagnosed infections and interrupting transmission chains^20,20,21,23^.

To address these challenges, public health programs have shifted their focus toward decentralizing testing and facilitating more direct access to the healthcare system. In France, alongside initiatives that expand community-based in 2024, direct access to STI testing has been extended through medical-biological laboratories (MBLs). This national reform allows individuals to present directly for a standardized panel of tests (HIV, HBV, syphilis, *Chlamydia*, and gonorrhea) without the need for a prior physician’s prescription. The reform was intended to broaden access, reduce administrative and psychological barriers, and enhance uptake among populations historically underserved by conventional physician-mediated pathways^24^. In addition to promoting STI detection in the general population, it is expected to particularly benefit groups with high incidence and transmission levels, such as males and young adults^4^.

Despite evidence supporting the benefits of public health policies that facilitate access to STI screening^24^, large-scale data evaluating the real-world impact of these measures remain limited. Leveraging a comprehensive dataset of STI tests performed in MBLs across most regions of France, this study aims to assess the reach and effectiveness of the 2024 national reform that introduced direct prescription-free access to STI screening through MBLs. Specifically, the study aims to characterize the demographic and behavioral profile of individuals undergoing STI testing according to their access modality, and to evaluate the probability of test positivity in relation to testing pathway, sex, and age group.

## METHODS

### Population

The data for this study were extracted from the Cerballiance health data warehouse that was authorized by the French authorities CNIL on March 18th, 2024 (authorization No: 2231450). This health data warehouse centralizes the biological test results from Cerballiance MBLs across most of metropolitan France, as well as from sites in La Reunion and Martinique. The following regions were excluded: Pays de la Loire, Bretagne and Martinique. All subjects whose data were included in this database were informed about the use of their data, and those who expressed opposition to the use of their data were excluded from the health database. This study included all subjects who attended Cerballiance MBLs for screening for *Chlamydia trachomatis*, *Neisseria gonorrhoeae*, HBV, syphilis, and HIV between March 1, 2025, and February 28, 2026. The first six months after policy implementation were excluded due to transitional coding and data-capture inconsistencies. This ensured that analyses relied only on stable, routine laboratory operations rather than early rollout effects, providing the most reliable and unbiased estimates. Patients with a documented history of hepatitis B vaccination were not eligible for HBV serology (HBsAg, HBsAb, HBcAb). For individuals reporting anal and/or oral sexual practices, additional anal and/or oropharyngeal swabs were collected as appropriate.

### Sexually transmitted infections screening protocol

Screening for STIs in the Cerballiance MBLs followed a standardized diagnosis protocol combining NAATs for bacterial agents (Hologic, Alinity, Seegene) and automated immunoassays for viral and bacterial infections (Alinity, Roche, Siemens). For *Chlamydia trachomatis* and *Neisseria gonorrhoeae*, detection was performed using NAATs, considered the gold standard for these pathogens. Testing included all relevant anatomical sites of exposure (urogenital specimen included first-void urine, vaginal swabs, rectal, and pharyngeal specimens) collected according to the individual’s sexual practices and risk profile (assessed by a clinician or self-reported). For viral and bacterial infections, including HIV, HBV, and syphilis, analyses were conducted on serum samples using fully automated immunoassay platforms. HIV screening was based on a combined antibody (Ab) and p24 antigen (Ag) assay. Confirmatory testing by immunoblot was performed at the Cerba laboratory. Results were interpreted according to the manufacturer’s instructions. HBV testing relied on detection of hepatitis B surface antigen (HBsAg) according to manufacturer instructions. Syphilis diagnosis was based on a treponemal-specific immunoassay for antibody detection, used for case identification within the screening panel. All tests were performed according to the manufacturers’ instructions and national quality assurance standards for diagnostic laboratories, ensuring reproducibility and comparability across sites. Tests yielding indeterminate or uninterpretable results were classified as inconclusive and excluded from analysis (0.2%). Positive cases were also categorized into mono-infections and co-infections (two or more positive STI detections per subject).

### Access modality and covariables

The screening access modality was documented by reception staff as prescription-based or prescription-free (PBA/PFA), harmonized across MBL sites, and integrated as a primary analytical variable. Additional covariates including age, sex, and region where the screening MBL was performed. For analytical clarity, age was categorized into the following groups: 15-19, 20-29, 30-39, 40-49, 50-59, 60-69, and ≥70 years.

### Statistical analysis

A descriptive and bivariate analysis was first conducted according to the STI screening access mode. Categorical variables were compared between groups using Pearson’s chi-squared test, and continuous variables using Student’s t-test. Differences in positivity rates and co-infection patterns were compared across access modalities, and an Upset plot was generated to visualize the distribution of coinfections within the study population.

Subsequently, a multivariable logistic regression model was fitted to evaluate the association between access modality (PBA *vs* PFA) and the probability of each positive STI results. The models were stratified by sex and adjusted for age groups, and MBL region (interaction sex*modality-access p<0.001). An interaction term between age group and access mode was included in each model, given its statistical significance and epidemiological relevance (*p*<0.05 in at least an interaction term in each model except syphilis and HIV models). Model results are presented as odds ratios (ORs) with 95% confidence intervals (95% CI). In addition, adjusted predicted probabilities of positivity were estimated for each age group according to access mode as an age-specific analysis to facilitate the interpretation. The Wald test was used to assess differences in predicted probabilities across age and access groups.

A logistic model with the same adjustment was fitted to evaluate the association between access mode and coinfection. For these models only individuals with positivity in at least one STI were included and coinfection was dichotomized as mono-infection vs co-infection (2 or more infections detected). All statistical analyses were performed using R (version 4.5.1), with data management and preprocessing conducted in Python 3.10.

## RESULTS

The total population screened of the Cerballiance MBLs during the first year of the facilitated STI screening program access included 1,462,792 subjects. Of this total, close to 3.1% were either under 15 years old or had missing data regarding sex or the region where the test was performed. Among the 1,416,640 remaining subjects, 1.4% showed inconsistent test results (*Chlamydia*-only or Gonorrhea-only test) or yielded uninterpretable results. Considering the total results, including each contact site as an individual test in the case of *Chlamydia and Gonorrhea*, close to 0.2% of these were uninterpretable and suggested re-evaluation. Finally, the study population consisted of 1,396,499 subjects, with 27.8% of them having undergone at least one STI screening at the MBLs using the PFA modality (Figure 1). During the campaign follow-up, the percentage of individuals who used PFA remained above 25.2% and 34.1% by month, with Mars 2025 being the month with the lowest proportion of individuals and February 2026, which was the month with the highest proportion of individuals (Fig S1).

**Figure 1.**
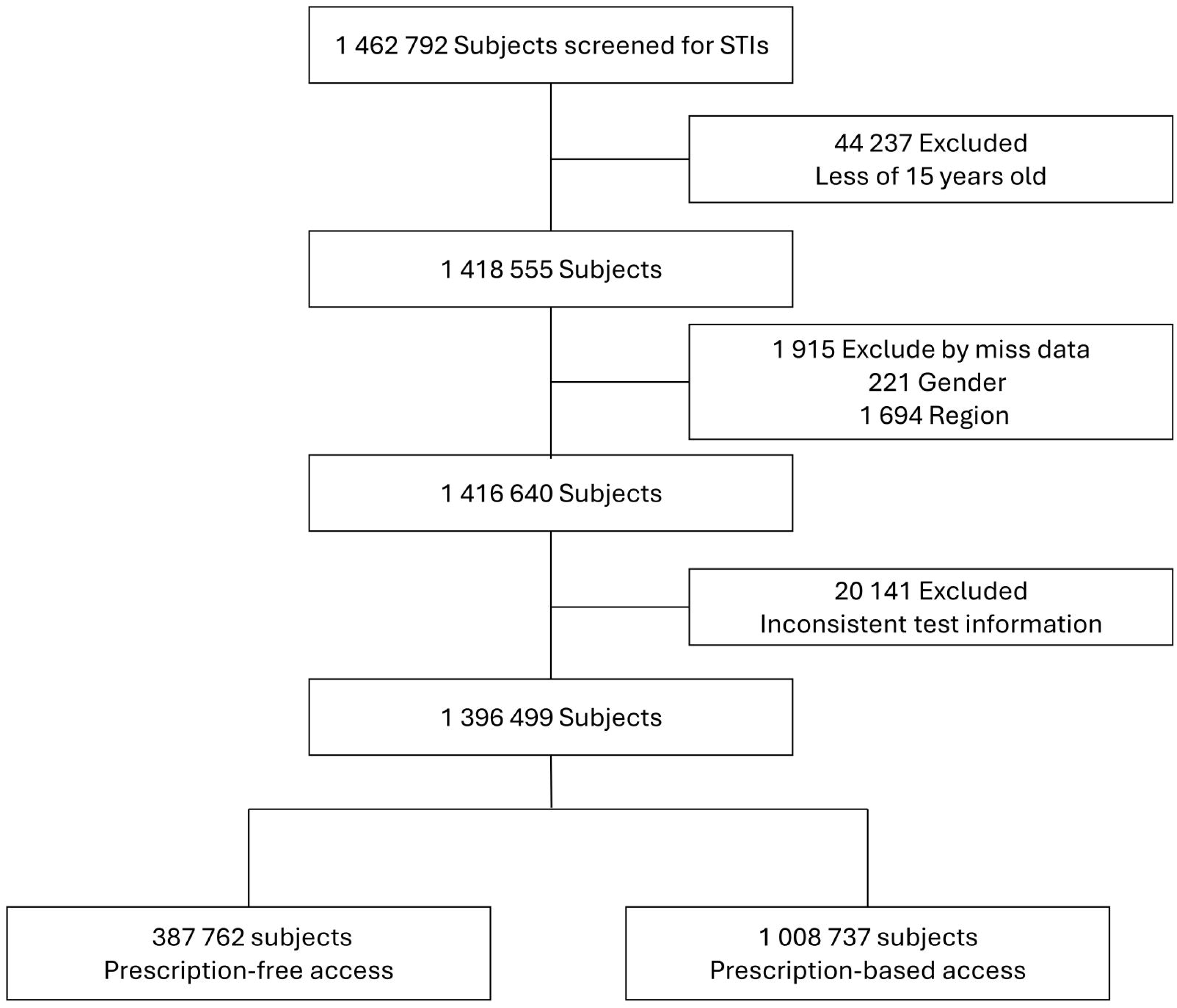
Study population within subjects screened for STIs. Subjects included from Mars 2025 until Mars 2026. STIs: Sexually transmitted infections.

Table 1 presents the demographic and testing characteristics of individuals by access modality. Compared with those accessing testing through a prescription, individuals using the PFA modality were more frequently male than female (47.4% vs. 36.3%, p<000.1) and more often belonged to the 20–29 age groups (p<000.1). Testing patterns also differed between modalities. Individuals using PFA underwent a higher mean number of STI (mean=2.81, SD=1.37) compared with those using PBA (mean=2.53, SD=1.27; p < 0.001) and were more frequently screened for HIV (70.6% vs. 66.3%) as well as *Chlamydia* and gonorrhea (46.6% vs 38.2%, both *p*<0.001). In contrast, screening for HBV was more common among PBA (64.7% vs 62.8%, *p*<0.001). Positivity rates also between modalities. PFA users showed lower positivity rates for syphilis (3.4% vs. 1.2%), HBV (1.3% vs. 0.4%), and HIV (0.3% vs. 0.2%; all *p*<0.001). PBA users exhibited a higher positivity rate for *Chlamydia* (4.6% vs. 3.6%), whereas no differences were observed between for gonorrhea (1.4% for both populations, p=0.437). Proportion of co-infections was no different between both groups (p=0.715).

**Table 1.** Characteristics of analysis population in function of screening access modality.

| Characteristics | Prescription-based<br>(N=1,008,737) | Prescription-Free<br>(N=387,762) | Total<br>(N=1,396,499) | p value |
| --- | --- | --- | --- | --- |
| Sex |  |  |  |  |
| Female | 642690 (63.7%) | 203770 (52.6%) | 846460 (60.6%) | < 0.001 |
| Male | 366047 (36.3%) | 183992 (47.4%) | 550039 (39.4%) |  |
| Age (Years) |  |  |  |  |
| 15-19 | 52227 (5.2%) | 17672 (4.6%) | 69899 (5.0%) | < 0.001 |
| 20-29 | 276490 (27.4%) | 131705 (34.0%) | 408195 (29.2%) |  |
| 30-39 | 278825 (27.6%) | 74932 (19.3%) | 353757 (25.3%) |  |
| 40-49 | 149394 (14.8%) | 60189 (15.5%) | 209583 (15.0%) |  |
| 50-59 | 100508 (10.0%) | 50368 (13.0%) | 150876 (10.8%) |  |
| 60-69 | 71376 (7.1%) | 34138 (8.8%) | 105514 (7.6%) |  |
| 70+ | 79917 (7.9%) | 18758 (4.8%) | 98675 (7.1%) |  |
| Region |  |  |  |  |
| Auvergne-Rhône-Alpes | 83517 (8.3%) | 21953 (5.7%) | 105470 (7.6%) | < 0.001 |
| Bourgogne-Franche-Comté | 9598 (1.0%) | 2196 (0.6%) | 11794 (0.8%) |  |
| Centre-Val de Loire | 30414 (3.0%) | 6874 (1.8%) | 37288 (2.7%) |  |
| Hauts-de-France | 93797 (9.3%) | 25333 (6.5%) | 119130 (8.5%) |  |
| Île-de-France | 342098 (33.9%) | 116528 (30.1%) | 458626 (32.8%) |  |
| Normandie | 48428 (4.8%) | 20778 (5.4%) | 69206 (5.0%) |  |
| Nouvelle-Aquitaine | 141766 (14.1%) | 100337 (25.9%) | 242103 (17.3%) |  |
| Occitanie | 78699 (7.8%) | 57447 (14.8%) | 136146 (9.7%) |  |
| Provence-Alpes-Côte d'Azur | 129300 (12.8%) | 23805 (6.1%) | 153105 (11.0%) |  |
| Réunion | 51120 (5.1%) | 12511 (3.2%) | 63631 (4.6%) |  |
| STI Screening in whole population |  |  |  |  |
| Chlamydia | 385273 (38.2%) | 157349 (40.6%) | 542622 (38.9%) | < 0.001 |
| Gonorrhea | 385273 (38.2%) | 157349 (40.6%) | 542622 (38.9%) | < 0.001 |
| Syphilis (TSI) | 462346 (45.8%) | 258357 (66.6%) | 720703 (51.6%) | < 0.001 |
| HBV | 652461 (64.7%) | 243417 (62.8%) | 895878 (64.2%) | < 0.001 |
| HIV | 668392 (66.3%) | 273683 (70.6%) | 942075 (67.5%) | < 0.001 |
| Total IST screened | 2.53 (1.26) | 2.81 (1.37) | 2.61 (1.30) | < 0.001 |
| Positive Result in IST screened population |  |  |  |  |
| Chlamydia | 13931 (3.6%) | 7142 (4.6%) | 21073 (3.9%) | < 0.001 |
| Gonorrhea | 5316 (1.4%) | 2130 (1.4%) | 7446 (1.4%) | 0.437 |
| Syphilis (TSI) | 15762 (3.4%) | 3149 (1.2%) | 18911 (2.6%) | < 0.001 |
| HBV | 8266 (1.3%) | 996 (0.4%) | 9262 (1.0%) | < 0.001 |
| HIV | 2330 (0.3%) | 609 (0.2%) | 2939 (0.3%) | < 0.001 |
| Co-infection in IST positive population |  |  |  |  |
| Co-infection | 2866 (6.7%) | 871 (6.6%) | 3737 (6.7%) | 0.715 |
| Mono-infection | 39654 (93.3%) | 12229 (93.4%) | 51883 (93.3%) |  |

Mean and standard deviation in parentheses, unless otherwise indicated. STIs: Sexually transmitted infections; HIV: human immunodeficiency virus; HBV: hepatitis B virus; TSI: treponemal-specific immunoassay.

Figure 2 illustrates the distribution of all detected positive cases, detailing mono-infection, and co-infection patterns. *Chlamydia* was the most frequently identified mono-infection (n=18,669), followed by Syphilis detected by treponemal immunoassay (n=16,757), HBV (n=8,950), Gonorrhea (n=5,031) and HIV (n=2,476). The most common co-infection observed was the combination of *Chlamydia* and Gonorrhea (n=1356). These findings are further disaggregated by access modality in Figure S2, reinforcing the divergent positivity rates previously noted. Specifically, within the PBA group (Figure S2, Panel A), Syphilis detected by treponemal immunoassay emerged as the dominant mono-infection (n=13,989), reflecting both acute cases and persistent treponemal antibodies from past infections. *Chlamydia* was the second most common (n=12,158) mono-infection in PBA group. In contrast, among the PFA group (Figure S2, Panel B), *Chlamydia* was the most frequent mono-infection (n=6,511), followed by Syphilis (n=2,768). Despite these differences in the prevalence of single infections, the combination of *Chlamydia* and *Gonorrhea* remained the most common co-infection across both modalities, in both PBA (n=915) and PFA (n=441).

**Figure 2.**
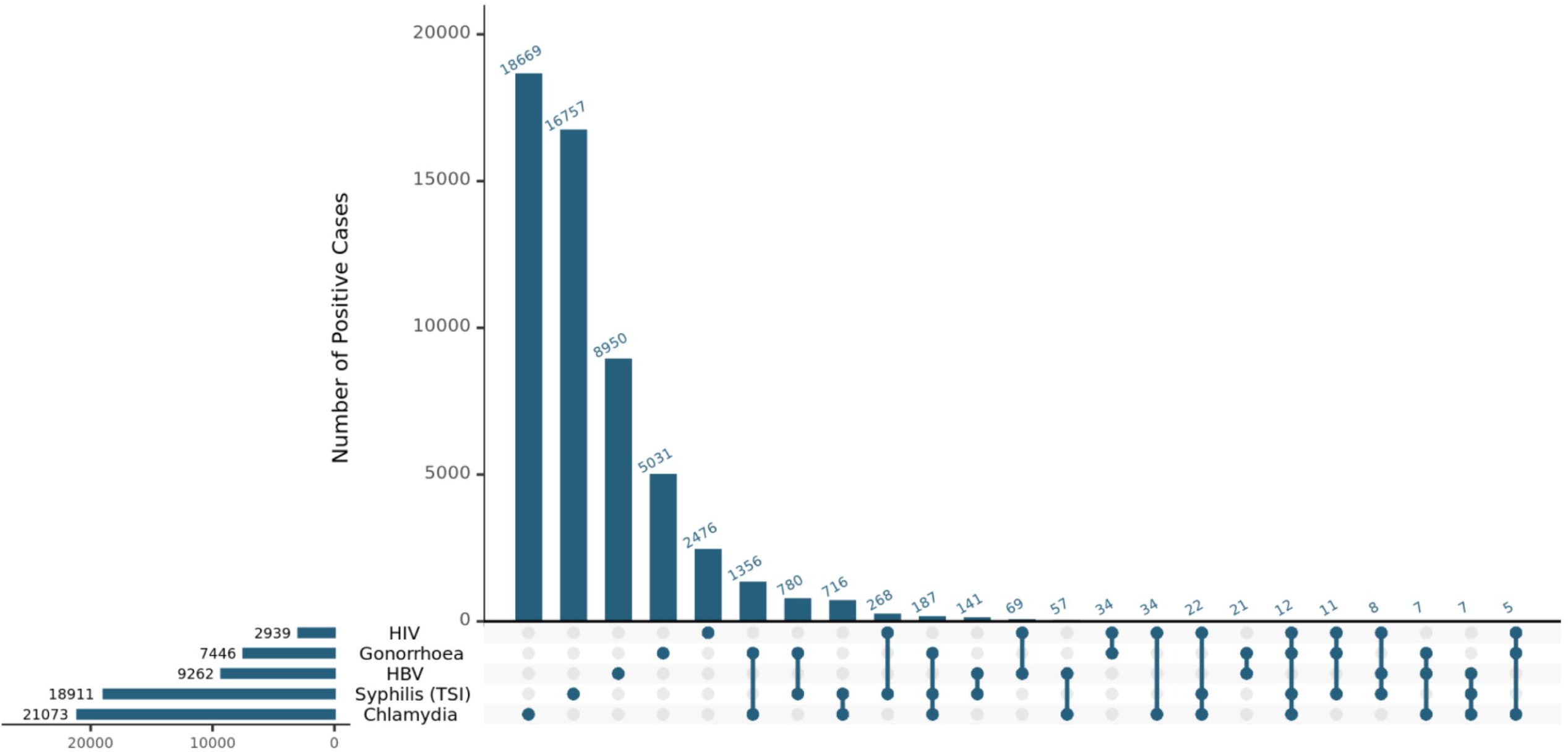
Distribution of positive cases of sexually transmitted infections and coinfection. HIV: human immunodeficiency virus; HBV: hepatitis B virus; TSI: treponemal-specific immunoassay.

Multivariable logistic models showed age-by-access interactions, indicating that the association between access modality and STI positivity varies significantly across different age groups (Figure 3; Table S1-S2). Among men, PFA showed no significant overall association with *Chlamydia* positivity (OR 0.92; 95% CI 0.79–1.08). However, significant age-specific reductions in positivity were observed through interaction effects. Compared with men aged 15–20 years, the effect of PFA was associated with progressively lower odds of *Chlamydia* infection from age 30 onward, with ORs of 0.71 (30–39 years), 0.51 (40–49 years), 0.42 (50–59 years), 0.28 (60–69 years), and 0.27 (70+ years), all p < 0.01. Among women, PFA was associated with higher overall odds of *Chlamydia* positivity (OR 1.23; 95% CI 1.10–1.38). Interaction terms indicated that this increase was concentrated in younger age groups: OR 1.22 for ages 20–29 (p = 0.002), while no significant associations were observed in older groups. Age-specific predicted probabilities allow observed clearly that among men, PFA produced virtually no change at ages 20–29 (7.1% under PBA vs. 7.0% under PFA) but was associated with lower predicted positivity in older men (4.2% vs. 2.8% at ages 30–39 and 3.4% vs. 1.6% at ages 40–49). In contrast, women showed higher predicted positivity under PFA in younger age groups, rising from 6.9% to 8.3% at ages 15–19 and from 4.4% to 6.4% at ages 20–29, while changes in older women were less pronounced (Figure 2).

**Figure 3.**
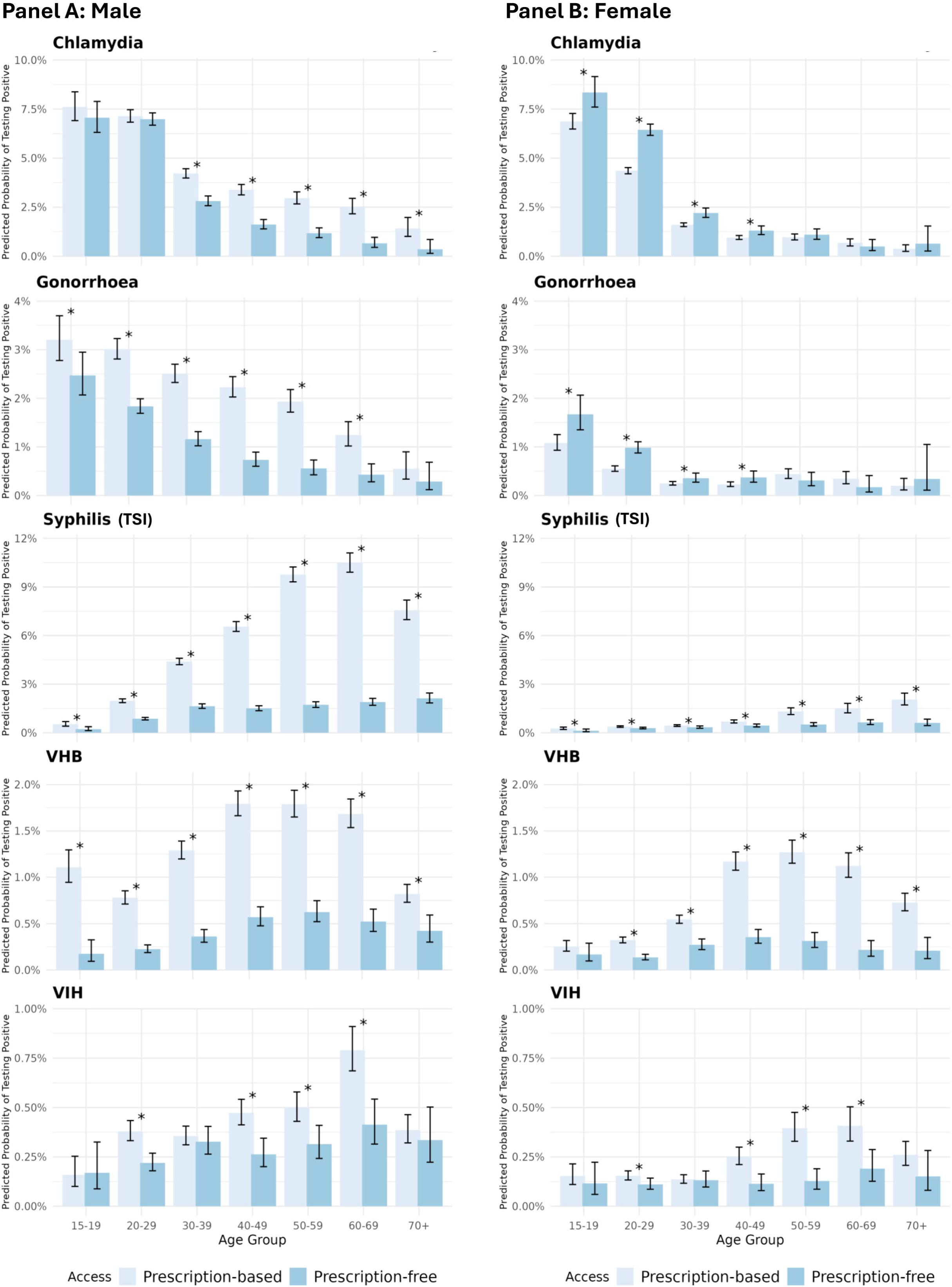
Predicted probabilities of sexually transmitted infection positivity by access modality. Logistic model adjusted to region and with interaction between age and access modality. Details of prediction in Table S2. HIV: human immunodeficiency virus; HBV: hepatitis B virus; TSI: treponemal-specific immunoassay.

**Figure 4.**
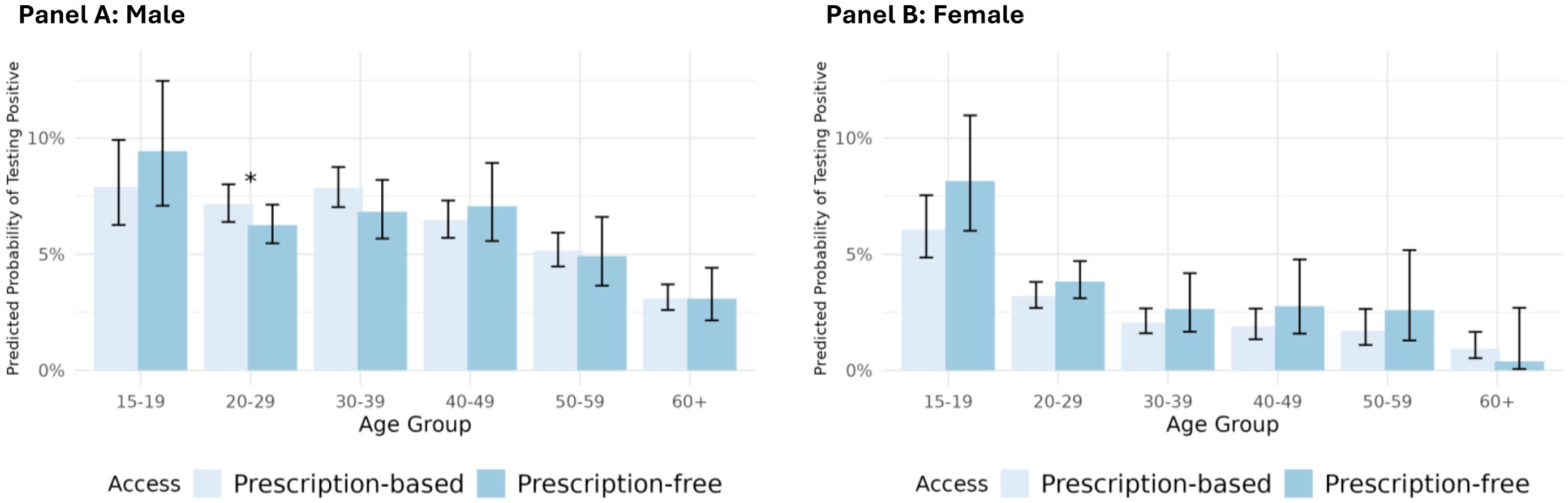
Probabilities of sexually transmitted co-infection detection by access modality in population where infection has already been detected. Logistic model adjusted to region and with interaction between age and access modality.

For gonorrhea, PFA was associated with lower overall positivity in men (OR=0.76; 95%CI=0.61-0.96) and higher positivity in women (OR=1.56; 95%CI=1.21-1.99). Among men, age-specific interaction terms showed that the reduction in positivity under PFA became more pronounced at older ages, with significantly lower odds at 30-39 years (OR=0.60; p<0.001), 40-49 years (OR=0.42; p<0.001), 50-59 years (OR=0.37; p<0.001), and 60-69 years (OR=0.45; p=0.002), while no differences were observed at younger ages. In contrast, women exhibited consistently higher odds of gonorrhea positivity under PFA, with no interaction effects indicating variation by age group. Age-specific predicted probabilities made this pattern more apparent: among adolescents (15-19 years), predicted positivity under PFA decreased from 3.2% to 2.5% in men but increased from 1.1% to 1.7% in women. Similar divergence was observed at ages 20-29, where men showed lower predicted positivity under PFA (3.0% to 1.8%), while women showed higher predicted positivity (0.6% to 1.0%). Across older ages, predicted positivity continued to decline in men under PFA, whereas changes among women remained small.

For syphilis, PFA was associated with lower overall odds of treponemal-test positivity in both men (O=0.42; 95%CI=0.23-0.73) and women (OR=0.46; 95%CI=0.22-0.86). Age-interaction terms in men indicated that these differences were more pronounced at specific ages, including 40-49 years (OR 0.51; p = 0.025), 50-59 years (OR=0.38; p=0.001), and 60–69 years (OR=0.39; p=0.002), while in women none of the interactions reached significance (all p>0.14). Age-specific predicted probabilities reflected this pattern: in men, treponemal-test positivity was systematically lower under PFA across all ages, from 6.6% to 1.5% at ages 40-49 and from 10.5% to 1.9% at ages 60-69; whereas differences in women were small in absolute terms and remained within very low predicted probability levels (all <2%).

For hepatitis B surface antigen, the main effect for men indicated substantially lower odds of HBV detection under PFA (OR=0.16; 95%CI=0.08-0.28). Several interaction terms reached statistical significance, notably at ages 40-49 (OR=2.01; p=0.040), 50-59 (OR=2.20; p=0.020), and 70+ (OR=3.27; p=0.001). Although these coefficients, the predicted probabilities remained uniformly low across age groups. Predicted HBV positivity among men ranged from 0.8-1.8% under PBA and from 0.2-0.6% under PFA, with similar patterns across all age strata. Among women, neither the main effect (OR-0.66; 95%CI=0.35-1.14) nor the interaction terms reached statistical significance (all p>0.14). Predicted probabilities also showed lower HBV positivity under PFA at all ages, with 1.2% to 0.4% at ages 40-49 and 1.1% to 0.2% at ages 60-69. Overall, predicted positivity suggests systematically lower HBV detection under PFA in both sexes, despite formal statistical significance being limited to men.

For HIV, neither men (OR=1.06; 95%CI=0.45-2.31) nor women (OR=0.75; 95%CI=0.34-1.49) showed significant differences in detection between PFA and PBA. None of the age-interaction terms reached significance (all p>0.10), indicating no evidence of age-specific variation in the association. Predicted probabilities were uniformly low across all ages and displayed no consistent differences by access modality, matching the regression findings.

The co-infection model, restricted to the 55,620 individuals with at least one positive STI result, showed no significant association between PFA and co-infection in either men (OR=1.21; 95%CI=0.82-1.78) or women (OR=1.37; 94%CI=0.94-1.97). Age was the strongest predictor of co-infection, with steadily decreasing odds compared with the 15-19-year reference group (e.g., men aged 40-49: OR=0.80; women aged 40-49: OR=0.30; p<0.001). None of the age-interaction terms reached statistical significance in either sex (all p>0.10), indicating no evidence that the association between access modality and co-infection varied by age. Age-specific predicted probabilities confirmed that individuals aged of 15-19 had the highest co-infection burden, reaching 9.4% under PFA vs. 7.9% under PBA in men and 8.1% vs 6.1% in women. Predicted co-infection risk declined steadily with age in both sexes, and differences between PFA and PBA were small in absolute terms at older ages.

## DISCUSSION

This cross-sectional study of more than 1 million individuals showed that under the policy of facilitating access to STI testing without a prescription in MBLs, from 40,720 confirmed positive cases (*Chlamydia*, Gonorrhea, HBV and HIV) close to 26,7% were detected without a medical consultation. In addition, 3,149 individuals were detected with treponemal-positive syphilis under PFA, representing 16.6% of all 18,911 syphilis TSI-positive results. The findings of this study also enabled the observation that the men and individuals aged 20-39 constituted the most frequent users of PFA testing. Regarding the identification of positive cases under the PFA, it was observed that females were more likely to be diagnosed with *Chlamydia* and gonorrhea infections compared with women under PBA. In the male population, positive cases detected for Chlamydia infection were similar with both access modalities in the 15-29 age group. Consequently, syphilis, HBV, and HIV infections were detected in the population mainly through the conventional (prescription-based) pathway.

The substantial acceptance of the prescription-free modality among younger individuals (20-29 years old), particularly men, indicates that the reform effectively reduced structural and psychological barriers to testing^25–27^. Young adults frequently report cost concerns, stigma, and limited availability of clinical appointments as deterrents to STI screening, making streamlined, walk-in strategies especially beneficial^19,25,26,28,29^ Consequently, the demographic profile observed in this study is consistent with known barriers and reflects success in reaching populations that would not otherwise be tested^25,29^. Moreover, the identification of almost a quarter of all positive cases through PFA underscores the significant contribution of this modality to overall screening performance^26^. Consequently, direct access not only modifies the route of access for users but also adds diagnosis value by capturing individuals who might otherwise not be tested, as well as shortening the infectious period and possibly interrupting ongoing chains of transmission^25,26^.

A key observation of this study is the differential diagnosis profile between modalities: PFA was associated with higher detection of *Chlamydia* and gonorrhea, whereas HBV, syphilis, and HIV were more often diagnosed through conventional pathway. This pattern is consistent with epidemiological differences across pathogens where bacterial STIs are more prevalent among younger, highly sexually active populations^4,30–32^, while chronic viral infections and syphilis often involve populations more frequently engaged with medical care, including older adults and individuals in risk-specific follow-up programs^32–34^. These findings underscore the complementary roles of both access pathways^35,36^.

Our findings indicate no significant differences in coinfections detection between individuals tested under PFA and those tested under PBA. However, PFA use was consistently more frequent among women across all age groups. This pattern may reflect differences in healthcare-seeking behavior, with women more often pursuing seek testing in response to symptoms, partner notification, or perceived exposure, which can increase the likelihood of identifying available testing modalities and a tendency to select the most rapid or convenient access pathway^37,38^. In contrast, the male population using prescription-free access appears more heterogeneous, possibly including a larger proportion of individuals at lower baseline risk.

The presence of age-related trends for HBV, syphilis, and HIV detection in the prescription-based modality, but not in the prescription-free one, suggests that the conventional model remains critical for reaching older adults and individuals with established risk or chronic infections^24,35^. Prescription-free access therefore functions as an additive, rather than substitute, strategy: it broadens screening for bacterial STIs among younger populations while conventional medical pathways continue to serve groups at higher risk for viral and chronic infections^24,36^.

This study, involving 720,703 individuals screened via treponemal immunoassay, identified 18,911 reactive cases. Model results confirm that these results primarily reflect the serological chronicity and cumulative disease burden across the lifespan rather than only acute transmission. These findings, showing predicted probabilities as high as 10.5% in older demographics, contrast with the annual incidence reported by Santé Publique France, which typically ranges between 12 and 23 new diagnoses per 100,000 inhabitants for adult populations^39^. This divergence underscores that while national surveillance captures annual diagnostic flows, our data reveals the extensive historical reservoir of lifetime exposure. In case of positive ELISA test, a reflex VDRL is added, which allows to differentiate past and current syphilis infection. This data will be available in further study. Furthermore, the significantly lower likelihood of positivity in the prescription-free cohort suggests that while direct access through biomedical laboratories successfully captures a younger demographic, the medicalized pathway remains the primary detector of the population’s chronic serological stock.

This study was conducted on a large population, providing substantial statistical power to detect age-and sex-specific associations that would have been difficult to observe in smaller samples. The broad geographic distribution of the individuals included also enhanced the interpretability of the findings by incorporating heterogeneity across multiple regions. In contrast, when compared to the PREVIST^40^ study, the positive rates obtained were more important in the present study. In fact, PREVIST study showed a prevalence for *Chlamydia* of 0.58% for men and 0.93% for women, highest among those aged 25-29. Differences between both studies should be explained by the fact that our study population is not representative of the general French population, as it includes individuals who had at higher risk because they mentioned all sexual exposure in the questionnaire. Additionally, screening in the PREVIST study was performed at a single anatomical site (vaginal or urine samples), whereas in our study, samples were possibly collected from one, two or three anatomical sites oral, genital, and anal according to sexual practice), which likely increased the detection rate and contributed to the higher positivity observed. However, these results must be considered with several limitations. First, although the dataset included individuals from various parts of France, some major regions, such as Grand-Est and Bretagne, were not represented, which may slightly limit the generalizability of the prevalence to the national population. Second, the study population consisted primarily of individuals who accessed services within a specific network of MBLs. This may introduce selection bias, as these users are likely already engaged in a structured healthcare pathway and may not fully reflect the general population, particularly younger individuals who are often less connected to formal care systems. Third, the absence of additional demographic variables such as educational level, socioeconomic status, health behaviors, and comorbidities prevented a more comprehensive sociodemographic characterization of individuals using the prescription-free modality. We anticipate enriching our database in the future to better quantify these dimensions and evaluate their influence on test utilization and outcomes. Fourth, co-infection modeling relied on a dichotomous variable, although we acknowledge that co-infection patterns may involve more complex interactions that future research should explore using more granular analytical frameworks.

### Conclusion

Our findings suggest that the French public health programs facilitating more direct access to STI testing, whereby testing is now available without a medical prescription, has resulted in enhanced early detection within high-transmission groups when compared with the previous system of conventional access. Moreover, it emphasizes the ongoing necessity for clinical pathways for conditions necessitating a more comprehensive evaluation, such as HIV, HBV, and mainly syphilis. Thus, the integration of flexible, easily accessible testing strategies as an effective component of national STI control efforts is supported by the results of this study.

## Abbreviations

STI: Sexually Transmitted Infection
NAATs: Nucleic Acid Amplification Tests
MBL: Medical Biological Laboratory
HIV: Human Infection Virus;
HBV: Hepatitis B virus
TSI: treponemal-specific immunoassay.

## Acknowledgements

We extend our sincere gratitude to all users in the Cerballiance laboratory network who consented to the use of their data, making this study possible. We thank STI study group and the laboratory teams whose work in data collection and quality assurance ensured the reliability of the dataset. Finally, we acknowledge the Data and IT teams at Cerba HealthCare Gestion for providing the logistical and technical resources necessary for the secure handling and processing of the data.

## STI study group

Benoit Chassaing^4^, Philippe Clech^8^, Jean Philippe Galhaud^7^, Lucie Messeant^10^, Sylvain Metge^3^, Pierre Bancons^13^, Leonard Rizzi^14^, Jean-Christophe Roig^6^, Sabine Trombert^12^, Mathilde Roussel^12^, Jean-Marc Aubert^1^.

## Contribution

Conceptualization: SHB, VG, SA, AD, SR, LP, NL, EC, AD, ELN, OB, ACS, CF, RL, CF, SL, BV, LE, CL; Data curation, Formal analysis, Methodology, Visualization: AGS, SHB, CV, DF; Funding acquisition: SHB, CL, CV; Investigation: SHB, VG, BG; Project administration, Resources, Software: SHB, CV CL; Supervision: SHB; Writing-original draft: AGS, SHB, VG, BG; Writing – review & editing, Validation: SA, AD, SR, LP, NL, EC, AD, ELN, OB, ACS, CF, RL, CF, SL, BV, LE, CL .

## Data availability

All data presented in the manuscript or in the supplementary material may be requested from the Cerba Healthcare Data Team upon submission of a justified request.

## Ethical statement

Data for this study were extracted from the Cerballiance Health Data Warehouse, authorized by the French Data Protection Authority (CNIL) on March 18, 2024 (authorization No. 2231450). Individuals who objected to the use of their data or who could be directly contacted for consent were excluded in accordance with CNIL regulations. The general results of this study will be communicated within Cerballiance laboratories to ensure that users are informed about the use of their data.

## REFERENCES

1. Elendu, C. et al. Global perspectives on the burden of sexually transmitted diseases: A narrative review. Medicine (Baltimore) 103, e38199 (2024).

2. Deng, M. et al. Trends in the incidence of common sexually transmitted infections at the global, regional and national levels, 1990–2021: results of the Global Burden of Disease 2021 study. Trop. Med. Health 53, 70 (2025).

3. Newman, L., et al. Global Estimates of the Prevalence and Incidence of Four Curable Sexually Transmitted Infections in 2012 Based on Systematic Review and Global Reporting. PloS One 10, e0143304 (2015).

4. Sexually transmitted infections (STIs). https://www.who.int/news-room/fact-sheets/detail/sexually-transmitted-infections-(stis).

5. Maqsood, N., Daniel, J. & Forsyth, S. The Risk of Pelvic Inflammatory Disease in Women Infected With Chlamydia (Chlamydia trachomatis): A Literature Review. Cureus 16, e66316 (2024).

6. Wiesenfeld, H. C., Hillier, S. L., Meyn, L. A., Amortegui, A. J. & Sweet, R. L. Subclinical pelvic inflammatory disease and infertility. Obstet. Gynecol. 120, 37–43 (2012).

7. Liu, L. et al. Chlamydia infection, PID, and infertility: further evidence from a case-control study in China. BMC Womens Health 22, 294 (2022).

8. Smolarczyk, K. et al. The Impact of Selected Bacterial Sexually Transmitted Diseases on Pregnancy and Female Fertility. Int. J. Mol. Sci. 22, 2170 (2021).

9. Berhil, T., Radi, F. Z., El Boussaadani, B. & Raissouni, Z. Tertiary syphilis and cardiovascular disease: the united triad: case report. Eur. Heart J. Case Rep. 8, ytae013 (2024).

10. Machado, M. de N., Trindade, P. F., Miranda, R. C. & Maia, L. N. [Bilateral ostial coronary lesion in cardiovascular syphilis: case report]. Rev. Bras. Cir. Cardiovasc. Orgao Of. Soc. Bras. Cir. Cardiovasc. 23, 129–131 (2008).

11. Rizzo, G. E. M., Cabibbo, G. & Craxì, A. Hepatitis B Virus-Associated Hepatocellular Carcinoma. Viruses 14, 986 (2022).

12. Duberg, A.-S., Lybeck, C., Fält, A., Montgomery, S. & Aleman, S. Chronic hepatitis B virus infection and the risk of hepatocellular carcinoma by age and country of origin in people living in Sweden: A national register study. Hepatol. Commun. 6, 2418–2430 (2022).

13. Wong, N. S. Sexually transmitted infection test-and-treat. IJID Reg. 14, 100529 (2025).

14. Workowski, K. A. et al. Sexually Transmitted Infections Treatment Guidelines, 2021. MMWR Recomm. Rep. Morb. Mortal. Wkly. Rep. Recomm. Rep. 70, 1–187 (2021).

15. Ong, J. J. et al. Beyond behavioural change: prioritising structural solutions to control bacterial sexually transmitted infections. eClinicalMedicine 83, 103198 (2025).

16. Rönn, M. M., Mc Grath-Lone, L., Davies, B., Wilson, J. D. & Ward, H. Evaluation of the performance of nucleic acid amplification tests (NAATs) in detection of chlamydia and gonorrhoea infection in vaginal specimens relative to patient infection status: a systematic review. BMJ Open 9, e022510 (2019).

17. US Preventive Services Task Force et al. Screening for Chlamydia and Gonorrhea: US Preventive Services Task Force Recommendation Statement. JAMA 326, 949–956 (2021).

18. Martin, K., Wenlock, R., Roper, T., Butler, C. & Vera, J. H. Facilitators and barriers to point-of-care testing for sexually transmitted infections in low- and middle-income countries: a scoping review. BMC Infect. Dis. 22, 561 (2022).

19. Denison, H. J., Bromhead, C., Grainger, R., Dennison, E. M. & Jutel, A. Barriers to sexually transmitted infection testing in New Zealand: a qualitative study. Aust. N. Z. J. Public Health 41, 432–437 (2017).

20. Footman, A., Dagama, D., Smith, C. H. & Van Der Pol, B. A Systematic Review of New Approaches to Sexually Transmitted Infection Screening Framed in the Capability, Opportunity, Motivation, and Behavior Model of Implementation Science. Sex. Transm. Dis. 48, S58–S65 (2021).

21. Tilson, E. C. et al. Barriers to asymptomatic screening and other STD services for adolescents and young adults: focus group discussions. BMC Public Health 4, 21 (2004).

22. Ehlers, E., Kovaleski, L., Devaskar, S., Kennedy, S. & Plotzker, R. E. Facilitators and Barriers of Implementing Expanded Sexually Transmitted Infection Screening in California Family Planning Clinics. Sex. Transm. Dis. 52, 9–13 (2025).

23. Zucker, J. et al. Attitudes and Perceived Barriers to Sexually Transmitted Infection Screening Among Graduate Medical Trainees. Sex. Transm. Dis. 48, e149–e152 (2021).

24. Sumray, K., Lloyd, K. C., Estcourt, C. S., Burns, F. & Gibbs, J. Access to, usage and clinical outcomes of, online postal sexually transmitted infection services: a scoping review. Sex. Transm. Infect. 98, 528–535 (2022).

25. Reeves, J. M. et al. Exploring Facilitators and Barriers to STD/STI/HIV Self-Testing Among College Students in the United States: A Scoping Review. J. Prim. Care Community Health 15, 21501319241291758 (2024).

26. SPF. Facilitating the access to HIV testing at lower costs: ‘To the laboratory without prescription’ (ALSO), a pilot intervention to expand HIV testing through medical laboratories in France. https://www.santepubliquefrance.fr/import/facilitating-the-access-to-hiv-testing-at-lower-costs-to-the-laboratory-without-prescription-also-a-pilot-intervention-to-expand-hiv-testing.

27. Sonubi, T. et al. STI testing, diagnoses and online chlamydia self-sampling among young people during the first year of the COVID-19 pandemic in England. Int. J. STD AIDS 09564624231180641 (2023) doi:10.1177/09564624231180641.

28. Tilson, E. C. et al. Barriers to asymptomatic screening and other STD services for adolescents and young adults: focus group discussions. BMC Public Health 4, 21 (2004).

29. Alarcon, J. et al. Barriers to Testing for Sexually Transmitted Infections among HIV-Serodiscordant Couples: The Influence of Discrimination. Ethn. Dis. 30, 261–268.

30. National Academies of Sciences, E. et al. Patterns and Drivers of STIs in the United States. in Sexually Transmitted Infections: Adopting a Sexual Health Paradigm (National Academies Press (US), 2021).

31. Torrone, E. A. et al. Prevalence of sexually transmitted infections and bacterial vaginosis among women in sub-Saharan Africa: An individual participant data meta-analysis of 18 HIV prevention studies. PLOS Med. 15, e1002511 (2018).

32. Patel, C. G. & Tao, G. STI/HIV Testing and Prevalence of Gonorrhea and Chlamydia Among Persons with Their Specified-Type Sex Partner. Am. J. Med. 135, 196–201 (2022).

33. Turpin, R., Rosario, A. D. & Dyer, T. Barriers to syphilis testing among men who have sex with men: a systematic review of the literature. Sex. Health 17, 201–213 (2020).

34. Berry, S. A. et al. GONORRHEA AND CHLAMYDIA TESTING INCREASING BUT STILL LAGGING IN HIV CLINICS IN THE UNITED STATES. J. Acquir. Immune Defic. Syndr. 1999 70, 275–279 (2015).

35. Barnard, S. et al. Comparing the characteristics of users of an online service for STI self-sampling with clinic service users: a cross-sectional analysis. Sex. Transm. Infect. 94, 377–383 (2018).

36. Wilson, E., Leyrat, C., Baraitser, P. & Free, C. Does internet-accessed STI (e-STI) testing increase testing uptake for chlamydia and other STIs among a young population who have never tested? Secondary analyses of data from a randomised controlled trial. Sex. Transm. Infect. 95, 569–574 (2019).

37. Huckabay, L., Fisher, D. G., Reynolds, G. L., Rannalli, D. & Erlyana, E. Gender differences in risk taking behaviors for Chlamydia trachomatis. Health Care Women Int. 41, 1147–1165 (2020).

38. Prevalence of co-infections with other sexually… : International Journal of STD & AIDS. Ovid https://www.ovid.com/journals/ijstd/fulltext/10.1177/0956462419890496∼prevalence-of-co-infections-with-other-sexually-transmitted.

39. Amber Kunkel et al. Screening and diagnosis of HIV and three bacterial sexually transmitted infections among young people in france, 2014-2023. Bull. Épidémiologique Hebd. BEH **19–20**, 373–382 (2025).

40. Claire Sauvage, et al. Prévalence de l’infection à Chlamydia trachomatis, Neisseria gonorrhoeae et Mycoplasma genitalium chez les femmes et les hommes de 18-59 ans, en France hexagonale, enquête PrévIST. (2025).

