## Supplementary material for "Impact of prescription-free access to sexually transmitted infection screening tests in medical-biological laboratories: cross-sectional analysis of data from clinical laboratories in France": Suplemental Materials

**Supplemental materials**

**Figure S1. Temporal evolution of the number of subjects undergoing screening tests for sexually transmitted infections according to access modality.**

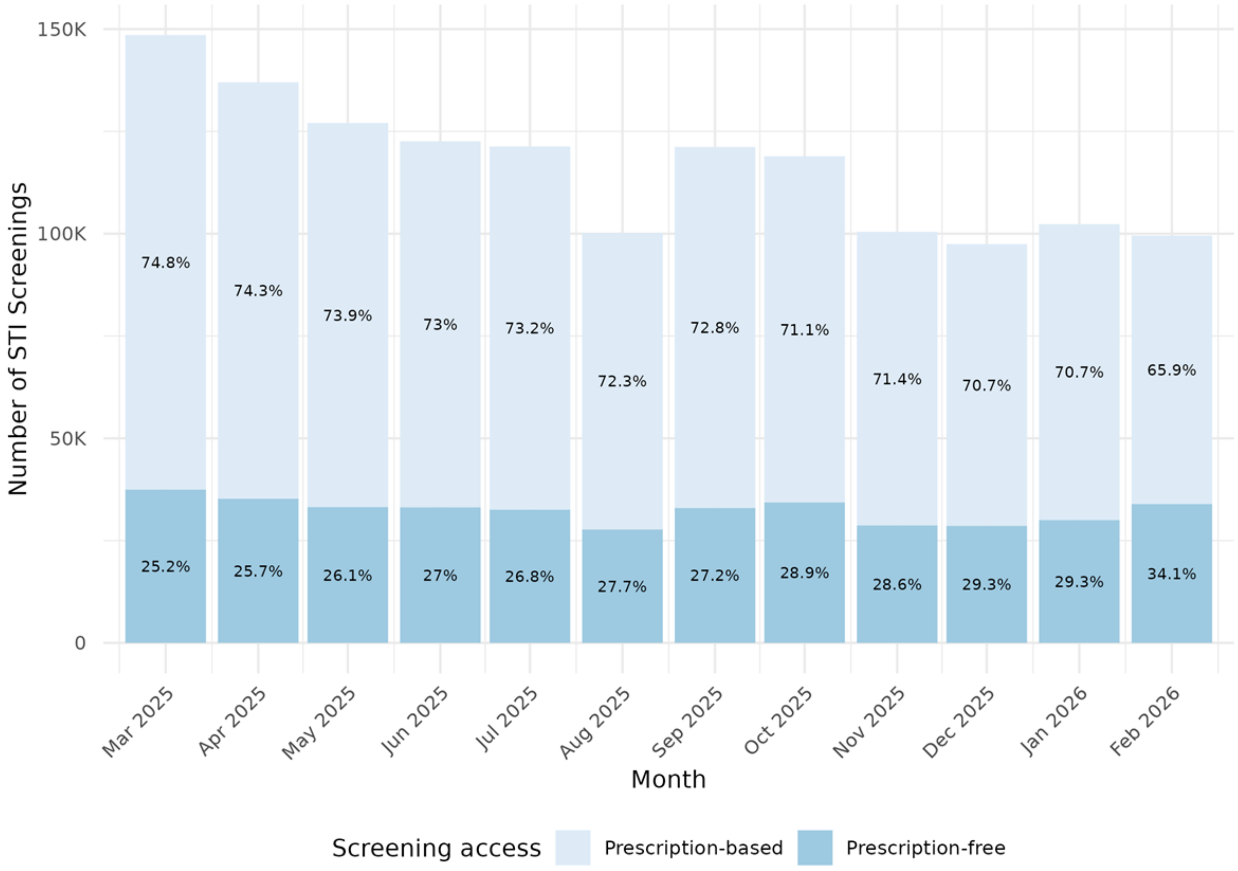


*STIs: Sexually transmitted infections.*

**Figure S2. Distribution of positive cases of sexually transmitted infections according to**

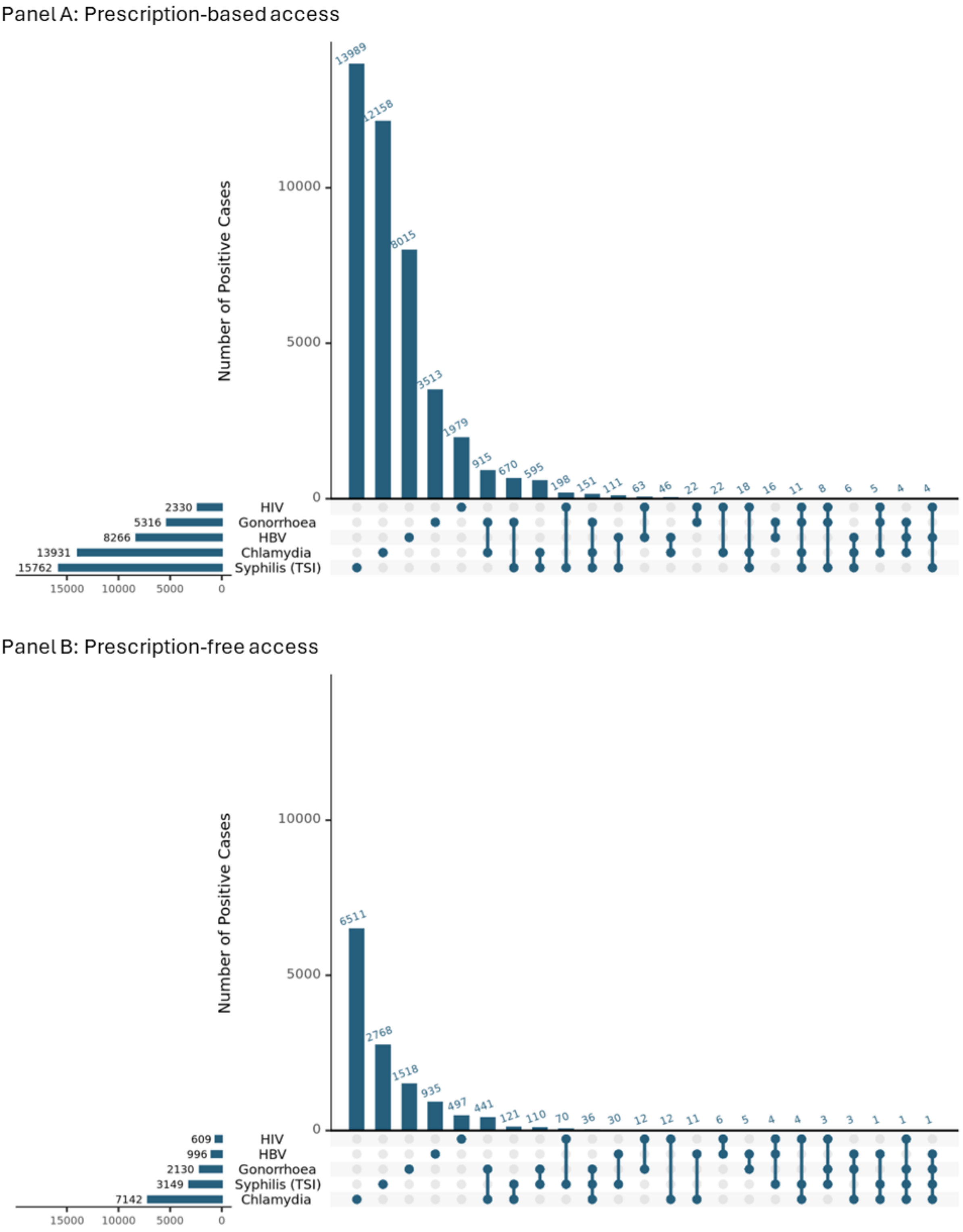


**access type**

*HIV: human immunodeficiency virus; HBV: hepatitis B virus; TSI: treponemal-specific immunoassay.*

**Table S1. Odds ratio of positive sexually transmitted infections. Multivariate logistic models result.**

| | Male | | | | Female | | | |
|---|---|---|---|---|---|---|---|---|
| **Chlamydia** | Odds Ratio | CI Lower | CI Upper | p Value | Odds Ratio | CI Lower | CI Upper | p Value |
| Access modality: Prescription-free | 0.92 | 0.79 | 1.08 | 0.304 | 1.23 | 1.10 | 1.38 | 0.000 |
| Age Group: 15-20 (Ref.) | | | | | | | | |
| 20-29 | 0.93 | 0.84 | 1.04 | 0.211 | 0.62 | 0.58 | 0.66 | 0.000 |
| 30-39 | 0.53 | 0.48 | 0.60 | 0.000 | 0.22 | 0.20 | 0.24 | 0.000 |
| 40-49 | 0.42 | 0.37 | 0.48 | 0.000 | 0.13 | 0.12 | 0.15 | 0.000 |
| 50-59 | 0.37 | 0.32 | 0.43 | 0.000 | 0.13 | 0.11 | 0.16 | 0.000 |
| 60-69 | 0.31 | 0.26 | 0.38 | 0.000 | 0.09 | 0.07 | 0.12 | 0.000 |
| 70+ | 0.17 | 0.12 | 0.24 | 0.000 | 0.05 | 0.03 | 0.08 | 0.000 |
| Region: Ile-de-France (Ref.) | | | | | | | | |
| Bourgogne-Franche-Comté | 0.56 | 0.51 | 0.61 | 0.000 | 0.65 | 0.60 | 0.69 | 0.000 |
| Centre-Val de Loire | 0.87 | 0.67 | 1.09 | 0.243 | 0.62 | 0.51 | 0.75 | 0.000 |
| Hauts-de-France | 1.24 | 1.10 | 1.38 | 0.000 | 1.15 | 1.04 | 1.26 | 0.005 |
| Île-de-France | 0.81 | 0.75 | 0.89 | 0.000 | 0.86 | 0.80 | 0.92 | 0.000 |
| Normandie | 1.11 | 0.99 | 1.23 | 0.065 | 0.70 | 0.61 | 0.81 | 0.000 |
| Nouvelle-Aquitaine | 0.98 | 0.92 | 1.04 | 0.561 | 0.86 | 0.81 | 0.91 | 0.000 |
| Occitanie | 0.96 | 0.88 | 1.04 | 0.305 | 0.83 | 0.77 | 0.89 | 0.000 |
| Provence-Alpes-Côte d'Azur | 0.81 | 0.74 | 0.88 | 0.000 | 0.95 | 0.88 | 1.03 | 0.259 |
| Réunion | 1.05 | 0.95 | 1.17 | 0.350 | 1.04 | 0.97 | 1.13 | 0.282 |
| Interactions | | | | | | | | |
| Prescription-free:20-29 | 1.06 | 0.90 | 1.25 | 0.494 | 1.22 | 1.08 | 1.39 | 0.002 |
| Prescription-free:30-39 | 0.71 | 0.59 | 0.86 | 0.000 | 1.12 | 0.95 | 1.33 | 0.180 |
| Prescription-free:40-49 | 0.51 | 0.40 | 0.64 | 0.000 | 1.11 | 0.88 | 1.40 | 0.354 |
| Prescription-free:50-59 | 0.42 | 0.32 | 0.56 | 0.000 | 0.92 | 0.67 | 1.24 | 0.571 |
| Prescription-free:60-69 | 0.28 | 0.18 | 0.43 | 0.000 | 0.59 | 0.31 | 1.06 | 0.094 |
| Prescription-free:70+ | 0.27 | 0.09 | 0.65 | 0.007 | 1.37 | 0.45 | 3.40 | 0.531 |

| **Gonorrhea** | Odds Ratio | CI Lower | CI Upper | p Value | Odds Ratio | CI Lower | CI Upper | p Value |
|---|---|---|---|---|---|---|---|---|
| Access modality: Prescription-free | 0.76 | 0.61 | 0.96 | 0.020 | 1.56 | 1.21 | 1.99 | 0.000 |
| Age Group: 15-20 (Ref.) | | | | | | | | |
| 20-29 | 0.94 | 0.81 | 1.09 | 0.397 | 0.51 | 0.43 | 0.59 | 0.000 |
| 30-39 | 0.78 | 0.67 | 0.91 | 0.001 | 0.23 | 0.19 | 0.27 | 0.000 |
| 40-49 | 0.69 | 0.59 | 0.81 | 0.000 | 0.21 | 0.16 | 0.26 | 0.000 |
| 50-59 | 0.60 | 0.50 | 0.71 | 0.000 | 0.40 | 0.31 | 0.52 | 0.000 |
| 60-69 | 0.38 | 0.30 | 0.48 | 0.000 | 0.32 | 0.21 | 0.46 | 0.000 |
| 70+ | 0.17 | 0.10 | 0.27 | 0.000 | 0.18 | 0.10 | 0.31 | 0.000 |
| Region: Ile-de-France (Ref.) | | | | | | | | |
| Bourgogne-Franche-Comté | 0.47 | 0.42 | 0.53 | 0.000 | 0.58 | 0.49 | 0.69 | 0.000 |
| Centre-Val de Loire | 0.46 | 0.30 | 0.68 | 0.000 | 0.40 | 0.22 | 0.67 | 0.001 |
| Hauts-de-France | 0.46 | 0.37 | 0.57 | 0.000 | 0.65 | 0.49 | 0.85 | 0.002 |
| Île-de-France | 0.56 | 0.50 | 0.63 | 0.000 | 0.69 | 0.58 | 0.82 | 0.000 |
| Normandie | 0.52 | 0.43 | 0.62 | 0.000 | 0.64 | 0.45 | 0.88 | 0.008 |
| Nouvelle-Aquitaine | 0.55 | 0.50 | 0.60 | 0.000 | 0.52 | 0.45 | 0.61 | 0.000 |
| Occitanie | 0.51 | 0.45 | 0.58 | 0.000 | 0.91 | 0.77 | 1.06 | 0.219 |
| Provence-Alpes-Côte d'Azur | 0.50 | 0.44 | 0.57 | 0.000 | 0.72 | 0.59 | 0.88 | 0.002 |
| Réunion | 0.74 | 0.64 | 0.85 | 0.000 | 1.13 | 0.96 | 1.33 | 0.133 |
| Interactions | | | | | | | | |
| Prescription-free:20-29 | 0.79 | 0.62 | 1.00 | 0.052 | 1.15 | 0.88 | 1.53 | 0.314 |
| Prescription-free:30-39 | 0.60 | 0.46 | 0.78 | 0.000 | 0.92 | 0.63 | 1.34 | 0.680 |
| Prescription-free:40-49 | 0.42 | 0.31 | 0.58 | 0.000 | 1.05 | 0.68 | 1.62 | 0.824 |
| Prescription-free:50-59 | 0.37 | 0.26 | 0.53 | 0.000 | 0.45 | 0.26 | 0.77 | 0.004 |
| Prescription-free:60-69 | 0.45 | 0.26 | 0.73 | 0.002 | 0.32 | 0.11 | 0.78 | 0.021 |
| Prescription-free:70+ | 0.68 | 0.22 | 1.79 | 0.459 | 1.10 | 0.25 | 3.57 | 0.888 |

*Continue...*

| | Male | | | | Female | | | |
|---|---|---|---|---|---|---|---|---|
| **Syphilis (TSI)** | Odds Ratio | CI Lower | CI Upper | p Value | Odds Ratio | CI Lower | CI Upper | p Value |
| Access modality: Prescription-free | 0.42 | 0.23 | 0.73 | 0.003 | 0.46 | 0.22 | 0.86 | 0.023 |
| Age Group: 15-20 (Ref.) | | | | | | | | |
| 20-29 | 3.80 | 2.94 | 5.02 | 0.000 | 1.43 | 1.10 | 1.89 | 0.010 |
| 30-39 | 8.70 | 6.74 | 11.47 | 0.000 | 1.65 | 1.27 | 2.18 | 0.000 |
| 40-49 | 13.27 | 10.28 | 17.51 | 0.000 | 2.59 | 1.98 | 3.46 | 0.000 |
| 50-59 | 20.48 | 15.86 | 27.05 | 0.000 | 4.99 | 3.76 | 6.72 | 0.000 |
| 60-69 | 22.18 | 17.12 | 29.35 | 0.000 | 5.68 | 4.18 | 7.82 | 0.000 |
| 70+ | 15.48 | 11.87 | 20.62 | 0.000 | 7.85 | 5.83 | 10.69 | 0.000 |
| Region: Ile-de-France (Ref.) | | | | | | | | |
| Bourgogne-Franche-Comté | 0.33 | 0.30 | 0.36 | 0.000 | 0.52 | 0.43 | 0.62 | 0.000 |
| Centre-Val de Loire | 0.35 | 0.26 | 0.44 | 0.000 | 0.22 | 0.09 | 0.45 | 0.000 |
| Hauts-de-France | 0.32 | 0.28 | 0.36 | 0.000 | 0.62 | 0.49 | 0.78 | 0.000 |
| Île-de-France | 0.28 | 0.25 | 0.30 | 0.000 | 0.29 | 0.24 | 0.36 | 0.000 |
| Normandie | 0.37 | 0.34 | 0.41 | 0.000 | 0.49 | 0.39 | 0.60 | 0.000 |
| Nouvelle-Aquitaine | 0.42 | 0.40 | 0.45 | 0.000 | 0.49 | 0.44 | 0.56 | 0.000 |
| Occitanie | 0.32 | 0.29 | 0.34 | 0.000 | 0.49 | 0.42 | 0.57 | 0.000 |
| Provence-Alpes-Côte d'Azur | 0.18 | 0.16 | 0.19 | 0.000 | 0.38 | 0.32 | 0.45 | 0.000 |
| Réunion | 0.29 | 0.27 | 0.32 | 0.000 | 1.49 | 1.32 | 1.68 | 0.000 |
| Interactions | | | | | | | | |
| Prescription-free:20-29 | 1.02 | 0.59 | 1.89 | 0.942 | 1.64 | 0.85 | 3.46 | 0.162 |
| Prescription-free:30-39 | 0.85 | 0.49 | 1.58 | 0.592 | 1.68 | 0.87 | 3.59 | 0.143 |
| Prescription-free:40-49 | 0.51 | 0.29 | 0.95 | 0.025 | 1.38 | 0.71 | 2.95 | 0.366 |
| Prescription-free:50-59 | 0.38 | 0.22 | 0.71 | 0.001 | 0.83 | 0.42 | 1.78 | 0.601 |
| Prescription-free:60-69 | 0.39 | 0.22 | 0.72 | 0.002 | 0.91 | 0.46 | 2.00 | 0.810 |
| Prescription-free:70+ | 0.63 | 0.35 | 1.17 | 0.122 | 0.63 | 0.31 | 1.41 | 0.235 |

| ***Hepatitis B Virus*** | Odds Ratio | CI Lower | CI Upper | p Value | Odds Ratio | CI Lower | CI Upper | p Value |
|---|---|---|---|---|---|---|---|---|
| Access modality: Prescription-free | 0.16 | 0.08 | 0.28 | 0.000 | 0.66 | 0.35 | 1.14 | 0.164 |
| Age Group: 15-20 (Ref.) | | | | | | | | |
| 20-29 | 0.70 | 0.59 | 0.84 | 0.000 | 1.27 | 1.01 | 1.62 | 0.047 |
| 30-39 | 1.17 | 0.99 | 1.38 | 0.065 | 2.15 | 1.72 | 2.73 | 0.000 |
| 40-49 | 1.63 | 1.39 | 1.93 | 0.000 | 4.65 | 3.71 | 5.90 | 0.000 |
| 50-59 | 1.63 | 1.38 | 1.93 | 0.000 | 5.05 | 4.01 | 6.45 | 0.000 |
| 60-69 | 1.53 | 1.29 | 1.83 | 0.000 | 4.46 | 3.51 | 5.74 | 0.000 |
| 70+ | 0.74 | 0.61 | 0.90 | 0.002 | 2.88 | 2.25 | 3.73 | 0.000 |
| Region: Ile-de-France (Ref.) | | | | | | | | |
| Bourgogne-Franche-Comté | 0.50 | 0.45 | 0.56 | 0.000 | 0.56 | 0.49 | 0.64 | 0.000 |
| Centre-Val de Loire | 0.56 | 0.39 | 0.78 | 0.001 | 0.49 | 0.32 | 0.71 | 0.000 |
| Hauts-de-France | 0.38 | 0.31 | 0.46 | 0.000 | 0.37 | 0.29 | 0.46 | 0.000 |
| Île-de-France | 0.25 | 0.21 | 0.29 | 0.000 | 0.30 | 0.25 | 0.35 | 0.000 |
| Normandie | 0.33 | 0.28 | 0.38 | 0.000 | 0.30 | 0.25 | 0.36 | 0.000 |
| Nouvelle-Aquitaine | 0.37 | 0.33 | 0.40 | 0.000 | 0.39 | 0.35 | 0.43 | 0.000 |
| Occitanie | 0.29 | 0.25 | 0.33 | 0.000 | 0.28 | 0.24 | 0.32 | 0.000 |
| Provence-Alpes-Côte d'Azur | 0.32 | 0.29 | 0.36 | 0.000 | 0.31 | 0.27 | 0.36 | 0.000 |
| Réunion | 0.33 | 0.28 | 0.39 | 0.000 | 0.44 | 0.38 | 0.52 | 0.000 |
| Interactions | | | | | | | | |
| Prescription-free:20-29 | 1.84 | 0.99 | 3.81 | 0.075 | 0.64 | 0.35 | 1.25 | 0.165 |
| Prescription-free:30-39 | 1.77 | 0.95 | 3.68 | 0.093 | 0.75 | 0.42 | 1.47 | 0.373 |
| Prescription-free:40-49 | 2.01 | 1.08 | 4.16 | 0.040 | 0.46 | 0.25 | 0.89 | 0.014 |
| Prescription-free:50-59 | 2.20 | 1.19 | 4.57 | 0.020 | 0.37 | 0.20 | 0.73 | 0.003 |
| Prescription-free:60-69 | 1.96 | 1.04 | 4.12 | 0.053 | 0.29 | 0.15 | 0.61 | 0.001 |
| Prescription-free:70+ | 3.27 | 1.63 | 7.15 | 0.001 | 0.43 | 0.19 | 0.97 | 0.038 |

*TSI: treponemal-specific immunoassay.*

*Continue...*

| | Male | | | | Female | | | |
|---|---|---|---|---|---|---|---|---|
| **Human Immunodeficiency Virus** | Odds Ratio | CI Lower | CI Upper | p Value | Odds Ratio | CI Lower | CI Upper | p Value |
| Access modality: Prescription-free | 1.06 | 0.45 | 2.31 | 0.883 | 0.75 | 0.34 | 1.49 | 0.445 |
| Age Group: 15-20 (Ref.) | | | | | | | | |
| 20-29 | 2.39 | 1.53 | 3.99 | 0.000 | 1.00 | 0.72 | 1.45 | 0.982 |
| 30-39 | 2.24 | 1.43 | 3.74 | 0.001 | 0.89 | 0.63 | 1.28 | 0.505 |
| 40-49 | 2.98 | 1.90 | 4.98 | 0.000 | 1.64 | 1.15 | 2.39 | 0.008 |
| 50-59 | 3.14 | 2.00 | 5.28 | 0.000 | 2.58 | 1.81 | 3.78 | 0.000 |
| 60-69 | 4.99 | 3.18 | 8.36 | 0.000 | 2.66 | 1.83 | 3.95 | 0.000 |
| 70+ | 2.43 | 1.52 | 4.12 | 0.000 | 1.70 | 1.15 | 2.55 | 0.008 |
| Region: Ile-de-France (Ref.) | | | | | | | | |
| Bourgogne-Franche-Comté | 0.49 | 0.39 | 0.62 | 0.000 | 0.60 | 0.45 | 0.77 | 0.000 |
| Centre-Val de Loire | 1.22 | 0.77 | 1.84 | 0.362 | 1.01 | 0.55 | 1.68 | 0.967 |
| Hauts-de-France | 0.69 | 0.50 | 0.92 | 0.015 | 0.70 | 0.48 | 0.97 | 0.041 |
| Île-de-France | 0.37 | 0.29 | 0.46 | 0.000 | 0.21 | 0.14 | 0.29 | 0.000 |
| Normandie | 0.51 | 0.39 | 0.65 | 0.000 | 0.54 | 0.40 | 0.72 | 0.000 |
| Nouvelle-Aquitaine | 0.48 | 0.41 | 0.55 | 0.000 | 0.50 | 0.41 | 0.60 | 0.000 |
| Occitanie | 0.55 | 0.45 | 0.66 | 0.000 | 0.78 | 0.64 | 0.95 | 0.016 |
| Provence-Alpes-Côte d'Azur | 0.78 | 0.67 | 0.89 | 0.001 | 0.75 | 0.62 | 0.89 | 0.002 |
| Réunion | 0.46 | 0.34 | 0.60 | 0.000 | 0.29 | 0.19 | 0.42 | 0.000 |
| Interactions | | | | | | | | |
| Prescription-free:20-29 | 0.54 | 0.24 | 1.31 | 0.151 | 0.95 | 0.45 | 2.20 | 0.905 |
| Prescription-free:30-39 | 0.87 | 0.38 | 2.08 | 0.735 | 1.29 | 0.60 | 3.01 | 0.537 |
| Prescription-free:40-49 | 0.52 | 0.23 | 1.28 | 0.135 | 0.60 | 0.27 | 1.44 | 0.228 |
| Prescription-free:50-59 | 0.59 | 0.26 | 1.45 | 0.229 | 0.43 | 0.19 | 1.04 | 0.049 |
| Prescription-free:60-69 | 0.49 | 0.21 | 1.20 | 0.102 | 0.62 | 0.27 | 1.53 | 0.275 |
| Prescription-free:70+ | 0.82 | 0.33 | 2.11 | 0.665 | 0.77 | 0.29 | 2.09 | 0.602 |

**Table S2. Probabilities of sexually transmitted infection positivity detection by access modality**

| | Male | | Female | |
|---|---|---|---|---|
| **Age** | **Prescription-based** | **Prescription-free** | **Prescription-based** | **Prescription-free** |
| *Chlamydia* | | | | |
| 15-19 | 0.076 (0.069 to 0.084) | 0.071 (0.063 to 0.079) | 0.069 (0.065 to 0.073) | 0.083 (0.076 to 0.092) |
| 20-29 | 0.071 (0.068 to 0.075) | 0.07 (0.067 to 0.073) | 0.044 (0.042 to 0.045) | 0.064 (0.062 to 0.067) |
| 30-39 | 0.042 (0.04 to 0.045) | 0.028 (0.026 to 0.031) | 0.016 (0.015 to 0.017) | 0.022 (0.02 to 0.025) |
| 40-49 | 0.034 (0.031 to 0.037) | 0.016 (0.014 to 0.019) | 0.01 (0.009 to 0.011) | 0.013 (0.011 to 0.015) |
| 50-59 | 0.03 (0.027 to 0.033) | 0.012 (0.01 to 0.014) | 0.01 (0.008 to 0.011) | 0.011 (0.009 to 0.014) |
| 60-69 | 0.025 (0.022 to 0.029) | 0.007 (0.005 to 0.01) | 0.007 (0.005 to 0.009) | 0.005 (0.003 to 0.009) |
| 70+ | 0.014 (0.01 to 0.02) | 0.004 (0.001 to 0.009) | 0.004 (0.002 to 0.006) | 0.006 (0.003 to 0.015) |
| *Gonorrhea* | | | | |
| 15-19 | 0.032 (0.028 to 0.037) | 0.025 (0.021 to 0.029) | 0.011 (0.009 to 0.013) | 0.017 (0.014 to 0.021) |
| 20-29 | 0.03 (0.028 to 0.032) | 0.018 (0.017 to 0.02) | 0.006 (0.005 to 0.006) | 0.01 (0.009 to 0.011) |
| 30-39 | 0.025 (0.023 to 0.027) | 0.012 (0.01 to 0.013) | 0.002 (0.002 to 0.003) | 0.004 (0.003 to 0.005) |
| 40-49 | 0.022 (0.02 to 0.024) | 0.007 (0.006 to 0.009) | 0.002 (0.002 to 0.003) | 0.004 (0.003 to 0.005) |
| 50-59 | 0.019 (0.017 to 0.022) | 0.006 (0.004 to 0.007) | 0.004 (0.004 to 0.005) | 0.003 (0.002 to 0.005) |
| 60-69 | 0.012 (0.01 to 0.015) | 0.004 (0.003 to 0.007) | 0.003 (0.002 to 0.005) | 0.002 (0.001 to 0.004) |
| 70+ | 0.006 (0.003 to 0.009) | 0.003 (0.001 to 0.007) | 0.002 (0.001 to 0.004) | 0.003 (0.001 to 0.011) |
| *Syphilis (TSI)* | | | | |
| 15-19 | 0.005 (0.004 to 0.007) | 0.002 (0.001 to 0.004) | 0.003 (0.002 to 0.003) | 0.001 (0.001 to 0.002) |
| 20-29 | 0.02 (0.019 to 0.021) | 0.009 (0.008 to 0.009) | 0.004 (0.003 to 0.004) | 0.003 (0.002 to 0.003) |
| 30-39 | 0.044 (0.042 to 0.046) | 0.016 (0.015 to 0.018) | 0.004 (0.004 to 0.005) | 0.003 (0.003 to 0.004) |
| 40-49 | 0.066 (0.063 to 0.069) | 0.015 (0.014 to 0.017) | 0.007 (0.006 to 0.008) | 0.004 (0.004 to 0.005) |
| 50-59 | 0.098 (0.093 to 0.102) | 0.017 (0.016 to 0.019) | 0.013 (0.011 to 0.015) | 0.005 (0.004 to 0.006) |
| 60-69 | 0.105 (0.099 to 0.111) | 0.019 (0.017 to 0.021) | 0.015 (0.012 to 0.018) | 0.006 (0.005 to 0.008) |
| 70+ | 0.076 (0.07 to 0.082) | 0.021 (0.018 to 0.024) | 0.02 (0.017 to 0.024) | 0.006 (0.004 to 0.008) |
| *Hepatitis B Virus* | | | | |
| 15-19 | 0.011 (0.009 to 0.013) | 0.002 (0.001 to 0.003) | 0.003 (0.002 to 0.003) | 0.002 (0.001 to 0.003) |
| 20-29 | 0.008 (0.007 to 0.009) | 0.002 (0.002 to 0.003) | 0.003 (0.003 to 0.004) | 0.001 (0.001 to 0.002) |
| 30-39 | 0.013 (0.012 to 0.014) | 0.004 (0.003 to 0.004) | 0.005 (0.005 to 0.006) | 0.003 (0.002 to 0.003) |
| 40-49 | 0.018 (0.017 to 0.019) | 0.006 (0.005 to 0.007) | 0.012 (0.011 to 0.013) | 0.004 (0.003 to 0.004) |
| 50-59 | 0.018 (0.017 to 0.019) | 0.006 (0.005 to 0.007) | 0.013 (0.012 to 0.014) | 0.003 (0.002 to 0.004) |
| 60-69 | 0.017 (0.015 to 0.018) | 0.005 (0.004 to 0.007) | 0.011 (0.01 to 0.013) | 0.002 (0.001 to 0.003) |
| 70+ | 0.008 (0.007 to 0.009) | 0.004 (0.003 to 0.006) | 0.007 (0.006 to 0.008) | 0.002 (0.001 to 0.004) |
| *Human Immunodeficiency Virus* | | | | |
| 15-19 | 0.002 (0.001 to 0.003) | 0.002 (0.001 to 0.003) | 0.002 (0.001 to 0.002) | 0.001 (0.001 to 0.002) |
| 20-29 | 0.004 (0.003 to 0.004) | 0.002 (0.002 to 0.003) | 0.002 (0.001 to 0.002) | 0.001 (0.001 to 0.001) |
| 30-39 | 0.004 (0.003 to 0.004) | 0.003 (0.003 to 0.004) | 0.001 (0.001 to 0.002) | 0.001 (0.001 to 0.002) |
| 40-49 | 0.005 (0.004 to 0.005) | 0.003 (0.002 to 0.003) | 0.003 (0.002 to 0.003) | 0.001 (0.001 to 0.002) |
| 50-59 | 0.005 (0.004 to 0.006) | 0.003 (0.002 to 0.004) | 0.004 (0.003 to 0.005) | 0.001 (0.001 to 0.002) |
| 60-69 | 0.008 (0.007 to 0.009) | 0.004 (0.003 to 0.005) | 0.004 (0.003 to 0.005) | 0.002 (0.001 to 0.003) |
| 70+ | 0.004 (0.003 to 0.005) | 0.003 (0.002 to 0.005) | 0.003 (0.002 to 0.003) | 0.002 (0.001 to 0.003) |

*Logistic model adjusted to region and with interaction between age and access modality. TSI: treponemal-specific immunoassay.*

**Table S3. Odds ratio of positive sexually transmitted co-infections. Multivariate logistic models result.**

| | Male | | | | Female | | | |
|---|---|---|---|---|---|---|---|---|
| **Co-infection Positive** | **Odds Ratio** | **CI Lower** | **CI Upper** | **p Value** | **Odds Ratio** | **CI Lower** | **CI Upper** | **p Value** |
| Access modality: Prescription-free | 1.21 | 0.82 | 1.78 | 0.320 | 1.38 | 0.94 | 1.98 | 0.089 |
| Age Group: 15-20 (Ref.) | | | | | | | | |
| 20-29 | 0.90 | 0.70 | 1.16 | 0.404 | 0.51 | 0.40 | 0.65 | 0.000 |
| 30-39 | 0.99 | 0.78 | 1.28 | 0.956 | 0.33 | 0.24 | 0.44 | 0.000 |
| 40-49 | 0.81 | 0.63 | 1.05 | 0.097 | 0.30 | 0.20 | 0.43 | 0.000 |
| 50-59 | 0.63 | 0.49 | 0.83 | 0.001 | 0.27 | 0.16 | 0.42 | 0.000 |
| 60+ | 0.37 | 0.28 | 0.50 | 0.000 | 0.15 | 0.08 | 0.26 | 0.000 |
| Region: Ile-de-France (Ref.) | | | | | | | | |
| Auvergne-Rhône-Alpes | 0.70 | 0.58 | 0.84 | 0.000 | 0.74 | 0.52 | 1.03 | 0.083 |
| Bourgogne-Franche-Comté | 0.32 | 0.13 | 0.62 | 0.003 | 0.67 | 0.20 | 1.60 | 0.434 |
| Centre-Val de Loire | 0.61 | 0.45 | 0.82 | 0.001 | 0.91 | 0.58 | 1.36 | 0.665 |
| Hauts-de-France | 0.62 | 0.50 | 0.76 | 0.000 | 0.92 | 0.68 | 1.24 | 0.613 |
| Normandie | 0.50 | 0.38 | 0.64 | 0.000 | 0.46 | 0.21 | 0.87 | 0.030 |
| Nouvelle-Aquitaine | 0.79 | 0.70 | 0.88 | 0.000 | 0.73 | 0.56 | 0.95 | 0.020 |
| Occitanie | 0.82 | 0.69 | 0.97 | 0.024 | 1.54 | 1.19 | 1.97 | 0.001 |
| Provence-Alpes-Côte d'Azur | 0.50 | 0.40 | 0.62 | 0.000 | 0.81 | 0.57 | 1.13 | 0.242 |
| Réunion | 0.56 | 0.44 | 0.71 | 0.000 | 1.47 | 1.12 | 1.89 | 0.004 |
| Interactions | | | | | | | | |
| Prescription-free:20-29 | 0.71 | 0.47 | 1.08 | 0.104 | 0.87 | 0.57 | 1.35 | 0.541 |
| Prescription-free:30-39 | 0.71 | 0.46 | 1.09 | 0.115 | 0.94 | 0.49 | 1.73 | 0.834 |
| Prescription-free:40-49 | 0.91 | 0.57 | 1.44 | 0.675 | 1.07 | 0.50 | 2.21 | 0.853 |
| Prescription-free:50-59 | 0.78 | 0.47 | 1.29 | 0.342 | 1.12 | 0.43 | 2.69 | 0.806 |
| Prescription-free:60+ | 0.82 | 0.47 | 1.41 | 0.477 | 0.30 | 0.02 | 1.61 | 0.255 |
